# Single-centre, non-randomised clinical trial at a tertiary care centre to investigate 1-year changes in social experiences and biomarkers of well-being after bariatric surgery in individuals with severe obesity: protocol for the Bariatric Surgery and Social Experiences (BaSES) study

**DOI:** 10.1101/2022.12.21.22283770

**Authors:** Daniela M. Pfabigan, Jens K. Hertel, Marius Svanevik, Morten Lindberg, Uta Sailer, Jøran Hjelmesæth

**Affiliations:** Department of Endocrinology, Obesity and Nutrition, Vestfold Hospital Trust, Tønsberg, Norway; Department of Behavioural Medicine, Institute of Basic Medical Sciences, Faculty of Medicine, University of Oslo, Oslo, Norway; Department of Biological and Medical Psychology, Faculty of Psychology, University of Bergen, Bergen, Norway; Department of Gastrointestinal Surgery, Vestfold Hospital Trust, Tønsberg, Norway; Department of Medical Biochemistry, Vestfold Hospital Trust, Tønsberg, Norway; Department of Endocrinology, Morbid Obesity and Preventive Medicine, Institute of Clinical Medicine, University of Oslo, Oslo, Norway

**Author notes:** Correspondence to: Daniela M. Pfabigan, Jonas Lies vei 91, 5009 Bergen, Norway M, F: N/A. US and JH contributed equally to this manuscript. Daniela M. Pfabigan, Jens K. Hertel, Marius Svanevik, Morten Lindberg, Uta Sailer, Jøran Hjelmesæth.

**Keywords:** social functioning, quality of life, loneliness, ghrelin, endocannabinoids

## Abstract

**Introduction:** Obesity is linked to increased loneliness and less enjoyment of social interactions. While bariatric surgery is the most effective treatment targeting severe obesity, there is limited understanding as to whether patients experience social interactions differently after surgery. The Bariatric Surgery and Social Experiences Study (BaSES) is designed to assess potential changes in how much patients enjoy and engage in daily social interactions 1 year after Roux-en-Y gastric bypass (RYGB) or sleeve gastrectomy (SG).

**Methods and analysis:** Single-centre, non-randomised clinical trial carried out at the Department of Endocrinology, Obesity and Nutrition at Vestfold Hospital Trust, Norway. Eligible patients (N=113) will undergo either RYGB, SG or single anastomosis sleeve ileal bypass (SASI). The primary outcome measure is change in the social experience score (assessed with a questionnaire) from a pre-surgery to a follow-up assessment 1 year after RYGB and SG. The respective changes after SASI will be assessed and considered exploratory.

**Ethics and dissemination:** The most recent protocol version of this study was reviewed and approved by the Regional Committee for Medical Research Ethics South East Norway (REK sør-øst A) on the 29^th^ of August 2022 (ref: 238406). The results will be disseminated to academic and health professional audiences and the public via publications in international peer-reviewed journals and conferences.

**Trial registration number:** NCT05207917 (ClinicalTrials.gov)

**Article Summary:** *Strengths and limitations of this study:*

- The primary outcome (frequency and quality of social interactions) is assessed over a 14-days period per measurement timepoint to comprehensibly capture daily variation.
- Multiple clinically relevant secondary outcomes including hair cortisol, endocannabinoid, and fasting ghrelin concentrations, cardiovascular risk factors and health-related and psychological patient-reported-outcomes are assessed.
- The sample size is limited and thus may not provide sufficient statistical power to compare the effects of RYGB and SG on secondary outcomes.

## INTRODUCTION

Obesity is one of the world’s most serious public health problems as defined by the World Health Organization. Currently, the most effective treatment at achieving lasting weight-loss and long-term reduction in obesity-related comorbidities is bariatric surgery (1,2). The two most commonly performed procedures are sleeve gastrectomy (SG) and Roux-en-Y gastric bypass (RYGB) (3), but more recently a combination of both (single anastomosis sleeve ileal bypass, SASI) has been introduced (4).

The effectiveness of bariatric surgery has most often been measured as weight loss and reductions in obesity-related complications and/or comorbidities, while effects on social interaction and subjective experience have received much less attention (see (5–7)). This appears to be an important omission as there is ample evidence that (supportive) social relationships promote health (8) and decrease mortality risk (9). Social relationships can affect a range of other health conditions such as cardiovascular disease, cancer, and immune function (8). However, individuals with obesity may experience social interactions as less positive than normal-weight individuals. Studies have reported that individuals with obesity avoid social events and relationships, but also career opportunities, shopping and other activities where they might feel observed (10–12) because of weight stigma. Such avoidance behaviour can lead to a “chronic disengagement” of diverse aspects of social life, which in turn might decrease interpersonal skills (13). Further investigations into the link between social behaviour and eating found that greater emotional eating was associated with greater social avoidance (14). Eating was described as a means to cope with loneliness on the one hand, while on the other hand aggravating feelings of being alone due to the stigma associated with obesity (15). As such, loneliness and obesity can create a vicious circle. Similarly, individuals with severe obesity report deriving less enjoyment from social contacts (16) and often feel more socially isolated than normal-weight individuals (17).

Only a few studies have investigated how bariatric surgery influences social interactions. One 10-year follow-up study found improvements in social interactions for bariatric surgery, but not for conventional weight loss treatment (18). A retrospective study observed a positive association between weight loss after bariatric surgery and the participants’ social connections (19). In qualitative studies, many participants reported that they received more positive social feedback following bariatric surgery (20), and also that they enjoyed social activities more than before (21), while others have described ambiguous feelings (22), or even negative psychosocial experiences (23).

The current clinical trial will address this important knowledge gap and investigate the effects of the two most common bariatric surgery procedures (RYGB and SG) on patients’ subjective experience of daily social interactions 1 year after surgery. Additionally, biological and psychological markers of social experiences will be assessed in this trial.

## Objectives

### Primary objectives

The primary objective of this study is to determine 1-year changes in patients’ subjective experience of daily social interactions after bariatric surgery (either RYGB or SG).

### Secondary objectives

Key secondary objectives are the examination of changes in variables assessing broader aspects of social experiences as secondary endpoints (affect and reactivity to social inclusion and exclusion, response to pleasant caress-like touch and one’s preferred velocity for self-applied caress-like touch) and biomarkers of well-being (cortisol and endocannabinoid concentrations from hair samples). Moreover, we will explore whether the primary and secondary endpoints also change in a short-term follow up (6 weeks after surgery; T1) and whether RYGB leads to larger changes in social interactions and in secondary endpoints related to social experiences than SG. In addition, we will explore changes in psychological (reward responsivity, social network and belonging, body image and interoceptive ability, self-reported eating behaviour) and health-related (obesity-related quality of life, psychological distress) patient reported outcomes (PROs), changes in gut hormones (ghrelin) and changes in anthropometric measures, body composition and cardiovascular risk factors in the surgery groups 6 weeks (T1) and 1 year (T2) after surgery. All outcomes will be considered exploratory.

### Trial design

This study is a single-centre, non-randomised clinical trial with two experimental groups (RYGB, SG). A third exploratory group will constitute of patients undergoing SASI.

## METHODS AND ANALYSIS

This protocol follows the SPIRIT reporting guidelines (24).

### Study setting

The study is conducted at a tertiary health-care centre, the Department of Endocrinology, Obesity and Nutrition, Vestfold Hospital Trust (Norway). Before the Covid-19 pandemic, about 180-200 bariatric surgeries were performed per year (about 65% RYGB and 35% SG). During the pandemic, the number of bariatric surgeries decreased to about 120 per year (with a similar distribution of the two surgery types as before the pandemic). The SASI procedure was first implemented in 2022.

### Patient and public involvement

A patient representative serves on the study steering committee to ensure that patients’ interests and opinions are taken into consideration during all study phases. The patients will be informed about published findings during the study period.

### Eligibility criteria

#### Inclusion criteria

- Eligibility for one of the surgery types
- Scheduled surgery
- Willingness and ability to give informed consent for study participation
- Aged between 18-80 years
- Good understanding of written and spoken Norwegian in order to answer the PRO-related questionnaires

#### Exclusion criteria

- Pregnancy and breast-feeding
- Severe chronic diseases such as endocrine, heart, neurological, lung, gastrointestinal or kidney conditions
- Cancer
- Acute psychotic episode

## Experimental groups

The current trial is not an intervention study. All participants will undergo one type of bariatric surgery and will thus be assigned to an experimental group, but this assignment will not directly influence study outcomes. The study outcome variables will only be observed before and after each surgery type. All surgical procedures will be performed laparoscopically by experienced surgical teams at Vestfold Hospital Trust.

### Experimental group 1: Roux-en-Y gastric bypass

In Roux-en-Y gastric bypass (RYGB), the left crus will be dissected free and significant hiatal hernias repaired with posterior cruraplasty. The minor curvature will be opened at the second vessel and the lesser sac entered. A 25 mL gastric pouch will be created by firing one horizontal and two vertical staple loads. The ligament of Treitz is then identified and a proximal loop of small intestine anastomosed to the pouch 60 cm from the ligament of Treitz with one linear stapler using the full length of the stapler, creating an antecolic, antegastric alimentary limb. The opening will then be closed using a single row, running absorbable suture. An entero- enteroanastomosis will be made 120 cm distal of the gastro-enteroanastomosis. The introductory opening is closed with a single row, running absorbable suture. Finally, the small intestine will be divided with one load between the gastro-entero-enteroanastomosis and the entero-enteroanastomosis in order to complete a bypass with an alimentary limb of 120 cm and a biliopancreatic limb of 60 cm.

### Experimental group 2: Sleeve gastrectomy

In sleeve gastrectomy (SG) a large part (80%) of the ventricle is removed. The greater curvature will be dissected free starting 1-2 cm from the pylorus up to the angle of Hiss. The left crus is then visualized and inspected for hiatal hernia. Clinically significant hiatal hernias will be repaired with posterior cruraplasty. The ventricle will then be lifted and any adhesions in the lesser sac divided. A 35 Fr bougie is placed down to the pylorus guiding the creation of a tubular sleeve with linear staplers. The first two loads are always purple, while tan loads are used for the rest of the ventricle. The last stapler is placed 5-10 mm laterally to the angle of Hiss. The staple line will then be inspected and secured with clips for additional haemostasis; no oversewing or buttressing material is routinely used.

### Experimental group 3: Single anastomosis sleeve ileal bypass

The sleeve gastrectomy is performed as described above, with the exception that the division of the stomach starts 6 cm proximal to the pylorus. The small bowel is measured 300 cm from the ileocecal valve, in sequences of 5 cm, with the small bowel stretched and markers placed on the graspers. The antrum is opened ventrally 5 cm proximal to the pylorus, just below the horizontal axis of canalis pylori and connected to the small bowel with a 30 mm stapled anastomosis completed with and absorbable running suture. The mesenteric defect is not closed.

### Concomitant care

Patients will follow standard treatment procedures at the Department of Endocrinology, Obesity and Nutrition, Vestfold Hospital Trust. There are no specific concomitant care or interventions that are permitted or prohibited during the trial.

### Outcome measures

#### Primary outcome measure

Change in social experience score (PRO: *The SOcial Experiences’ DaiLy Occurrence scale SOLO* (29)) from 4 weeks before surgery to 1 year after surgery. The SOLO is a short questionnaire assessing occurrence and quality of participants’ daily social interactions (both meaningful and superficial) in the 14 days following each visit.

#### Secondary outcome measures

Secondary outcome measures are listed in Box 1, and are assessed as summary measures such as mean/median or proportions (when appropriate) for each group. Outcome variables constitute the assessed values/scores per measurement time point and changes from baseline (Baseline: 4 weeks prior to surgery) to the short-term follow up (T1: 6 weeks after surgery) and to the long-term follow up (T2: 1 year after surgery); see Table 1.

##### Box 1: Secondary outcome measures

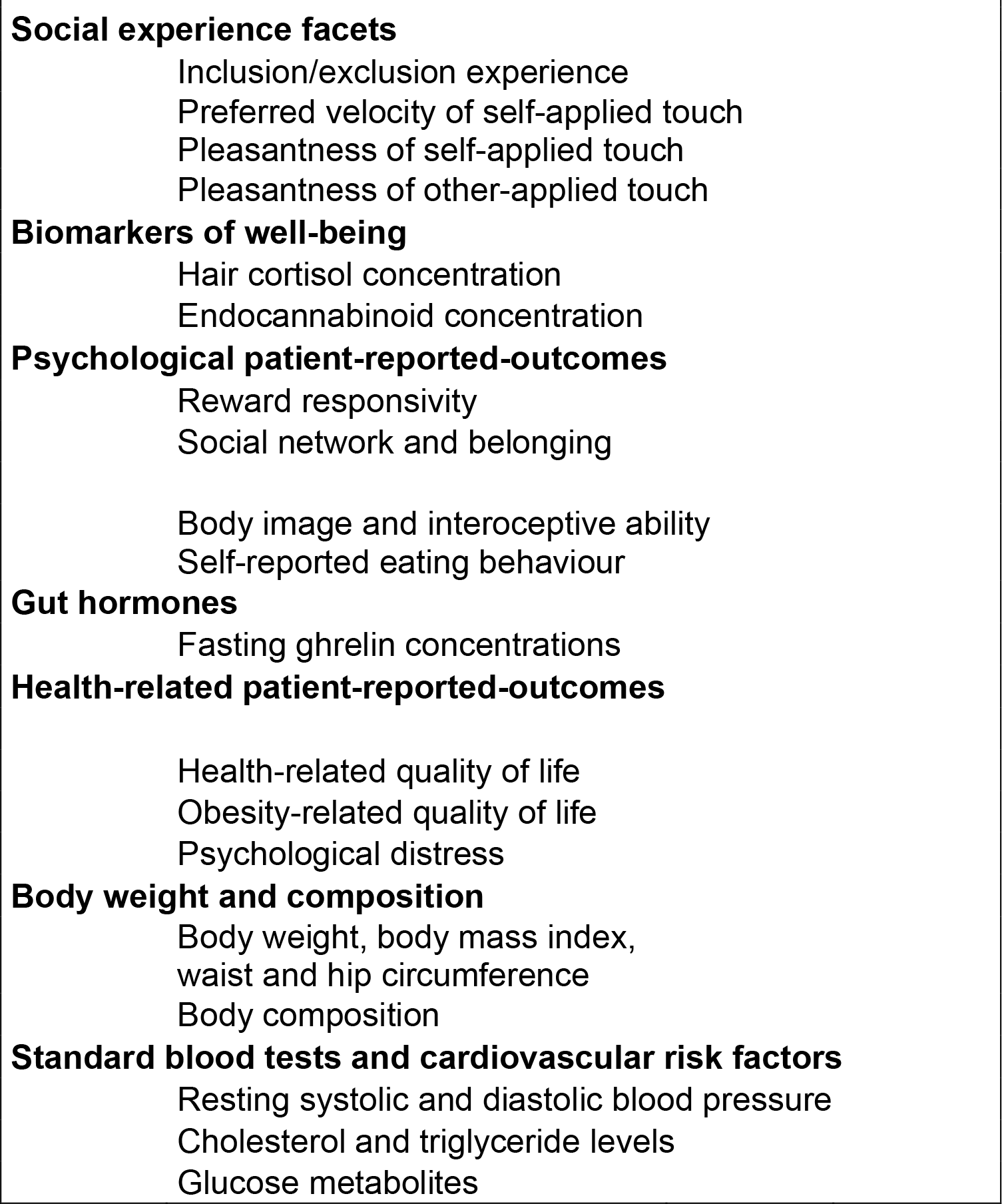

**Table 1.** Patient visit schedule

| Table 1. Patient visit schedule |  |  |  |  |  |  |  |
| --- | --- | --- | --- | --- | --- | --- | --- |
|  | Screening/<br>Information<br>meeting |  | Baseline |  | T1 Follow-up |  | T2 Follow-up |
|  |  |  | ~4 weeks<br>prior to<br>surgery |  | ~6 weeks<br>after<br>surgery |  | ~1 year<br>after<br>surgery |
| **VISIT** | **1** |  | **2** |  | **3** |  | **4** |
| Inclusion and exclusion criteria | x |  |  |  |  |  |  |
| Signed informed consent | x |  |  |  |  |  |  |
| Demographic data |  |  | x |  | x |  | x |
| PRO: SOLO |  |  | x |  | x |  | x |
| Psychological and health-related PROs |  |  | x |  | x |  | x |
| Social experience tasks |  |  | x |  | x |  | x |
| Regular medication |  |  | x |  | x |  | x |
| Hair samples |  |  | x |  | x |  | x |
| Blood samples |  |  | x |  | x |  | x |
| Body composition |  |  | x |  | x |  | x |
| Anthropometric measures |  |  | x |  | x |  | x |

### **Participant timeline** (Table 1 Patient visit schedule)

The BaSES study includes 4 time points where the patients are invited to the Department of Endocrinology, Obesity and Nutrition. Study enrolment started in May 2022 with screening meetings. The baseline assessment takes place approximately 4 weeks before surgery, the short-term follow-up is scheduled approximately 6 weeks after surgery (T1) and the long-term follow-up is scheduled approximately 1 year after surgery (T2). Up until the end of 2022, 26 patients underwent the baseline assessments. It is expected that the long-term follow-up (T2) will be finished in 2025.

### Sample size

This study has 1 primary outcome (“changes in the social experience score” from before to after surgery) that served as reference for sample size calculations. A small to medium effect size for this change from baseline to the long-term follow-up (T2) was assumed based on related literature (18,25–27). Power calculations were conducted with G*Power (28) for a dependent t-test with the following parameters: effect size of dz=0.4, two-tailed significance, alpha=.05, and a power of > 95%. The result recommended a sample size of 84 participants in total. Accounting for a drop-out rate of ≤ 35% (because of the time between baseline and the follow-up 1 year after surgery), we aim to recruit 113 participants in total, with similar patient numbers following RYGB and SG. Should the number of recruited patients in the two surgery groups differ considerably during inclusion, we will aim to recruit further participants in the smaller group to align the number of participants for both groups.

### Recruitment

Potential patients are informed about the BaSES study during courses and group sessions by trained staff at the Department of Endocrinology, Obesity and Nutrition. Patients from the waiting list for bariatric surgery will be individually contacted by phone and invited to an information/screening meeting in the weeks before surgery. Study inclusion will be assessed according to inclusion and exclusion criteria during this meeting. Patients who approve participation and pass eligibility criteria will then sign the informed consent form and will be enrolled in the study.

### Allocation – sequence generation and concealment

Participants will be numbered sequentially based on enrolment. No concealment regarding surgery type is implemented.

### Blinding

Due to the nature of the non-randomized controlled trial, neither participants nor staff will be blinded to the surgery type.

### Data collection methods

#### Primary outcome

Changes in the social experience score: After each BaSES study visit, participants are asked to fill in the *Social Experiences’ Daily Occurrence scale* (SOLO, (29)) in the 14 days following their visit. The SOLO is a 14-item questionnaire assessing occurrence and quality of one’s daily social interactions. The questionnaire is supposed to be filled in at the end of the day, taking between 2 and 5 minutes. It is available either in an online version or on paper. Participants will receive individual reminders each day in order to enhance compliance. A total item score will be calculated over the 14 assessment days at the 3 visits (Baseline, T1, T2). A change in the total SOLO score from baseline to the long-term follow-up (T2), irrespective of surgery type, constitutes the primary outcome.

#### Secondary and exploratory outcomes

See Supplementary Materials for a detailed description of the measurement of affect and reactivity to social inclusion and exclusion, and of subjective experience of caress-like other- and self-applied touch. Cortisol and endocannabinoid concentrations will be measured from hair samples and analysed by Dresden LabService GmBH (Dresden, Germany). The full list of the administered psychological and health-related questionnaires and a detailed description of the assessed anthropometric measures is provided in Supplementary Materials. Routine laboratory measurements will be performed at the Central Laboratory, Vestfold Hospital Trust, while C-peptide analyses will be performed at the Hormone Laboratory, Oslo University Hospital. Both laboratories are certified according to NO-EN ISO 15189. A detailed list of method principles, sample matric, units and analytical precision of biological outcomes are provided in Table S1 in Supplementary Materials.

#### Retention

Participants may withdraw from the study for any reason at any time but may also be excluded by study personnel in order to protect their safety and/or if they are unable/unwilling to comply with the study procedures. However, reasonable effort is made by the study personal to prevent attrition throughout the study period. Loss to follow-up measurement sessions of the primary outcome is estimated to be ≤ 35%.

### Data management

Trained study personnel will enter all data into online case report forms (CRF) during the study visits. CRFs will be saved on an encrypted server that requires two-factor authentication of authorized personal (TSD - Service for Sensitive Data, facilities, University of Oslo). SOLO data is directly transferred and saved on this server. The log files from the tasks assessing experience of social inclusion/exclusion and touch will be manually transferred to this server. Data integrity is continuously monitored by members of the steering committee.

Data will be stored in a pseudo-anonymised way because study-generated participant codes will be used. Participants’ data will be stored for a period of at least 5 years after completion of the study.

### Statistical methods

Descriptive data will be presented as mean (SD), median (range) or number (percentage). Within-group comparisons of differences between baseline and T2 (primary objective), and baseline and T1 will be calculated with dependent t-tests or non-parametric alternatives. Between-group comparisons of differences between RYGB and SG (and SASI in an explorative manner) will be calculated with independent t-tests, one-way ANOVAs or non-parametric alternatives. Interactions between within-group and between-group comparisons will be analysed using either a multilevel modelling approach (where appropriate; also to handle missing data) or mixed ANOVAs. Correlation and regression approaches will be used for the exploration of mediating or independent effects of surgery type, hormone concentrations or PRO scores on the primary outcome and selected secondary outcomes.

### Data monitoring

The steering committee consists of a team of healthcare professionals, researchers and a patient representative. Members of the steering committee meet every 12 months to safeguard the interests of patients participating in BaSES. It further monitors the progress and overall conduct of the clinical trial. Adverse events will be consecutively reported. The harms of the applied experimental assessments are extremely low and pose no risk for participants. Due to the low-risk nature of BaSES, the steering committee also assumes the role of the data monitoring committee.

## ETHICS AND DISSEMINATION

### Research ethics approval

The study protocol was registered in an international trial register (ClinicalTrials.gov). The study is conducted in accordance with Good Clinical Practice (GCP), International Council of Harmonization (ICH) guidelines and the latest revision of the Declaration of Helsinki.

### Protocol amendments

Significant amendments to the protocol have been and will be made only after ethical approval by the regional ethics committee.

### Informed consent

Individual informed consent was obtained in information/screening meetings consisting of either individual patients or small patient groups.

### Ancillary studies

Additional blood samples will be obtained and stored for use in future studies.

Information about storage and analyses of these samples is covered in the informed consent.

### Confidentiality

Each participant is given a study ID that will be used during data collection and analysis. The key linking participants’ names and study ID is stored on a high-security server with two- factor authentification. No participant information will be released outside the study.

### Access to data

Electronically authorized data access is available only to selected study personnel. Data analyses must either be performed according to the pre-planned statistical analysis plan or should be authorized by the study principal investigators before they are carried out. De-anonymized individual participant data can be made available following publication upon reasonable request to authors US and JH. Data will be shared according to the consent given by the participants and Norwegian laws and legislation.

### Ancillary and post-trial care

All patients will receive post-trial follow-up care according to national guidelines (30).

### Dissemination

The protocol and the results of the study will be published in international peer-reviewed journals in accordance with the ICMJE criteria for authorship (http://www.icmje.org/). Furthermore, study findings will be disseminated via scientific networks, conferences, professionals, policymakers and commissioners of weight management.

## Author contributions

Original idea: US, JH; Designed the study: US; DMP; JKH, JH; Study implementation: DMP; Provided input to methods: MS; ML; Data curation: DMP; Wrote the first version of the manuscript: DMP; Acquired funding: JH, US. All authors contributed and agreed to the final version of the manuscript.

## Patient consent for publication

Not required.

## Ethics approval

The most recent protocol version of this study was reviewed and approved by the Regional Committee for Medical Research Ethics South East Norway (REK sør-øst A) on the 29^th^ of August 2022 (ref: 238406).

## Data Availability

De-anonymized individual participant data can be made available following publication upon reasonable request to authors US and JH. Data will be shared according to the consent given by the participants and Norwegian laws and legislation.

## REFERENCES

1. Puzziferri N, Roshek TB, Mayo HG, Gallagher R, Belle SH, Livingston EH. Long-term follow-up after bariatric surgery: a systematic review. JAMA. 2014 Sep 3;312(9):934–42.

2. Jakobsen GS, Småstuen MC, Sandbu R, Nordstrand N, Hofsø D, Lindberg M, et al. Association of Bariatric Surgery vs Medical Obesity Treatment With Long-term Medical Complications and Obesity-Related Comorbidities. JAMA. 2018 Jan 16;319(3):291–301.

3. Angrisani L, Santonicola A, Iovino P, Vitiello A, Zundel N, Buchwald H, et al. Bariatric Surgery and Endoluminal Procedures: IFSO Worldwide Survey 2014. Obes Surg. 2017 Sep;27(9):2279– 89.

4. Sewefy AM, Atyia AM, Mohammed MM, Kayed TH, Hamza HM. Single anastomosis sleeve jejunal (SAS-J) bypass as a treatment for morbid obesity, technique and review of 1986 cases and 6 Years follow-up. Retrospective cohort. Int J Surg Lond Engl. 2022 Jun;102:106662.

5. Broadhead WE, Kaplan BH, James SA, Wagner EH, Schoenbach VJ, Grimson R, et al. The epidemiologic evidence for a relationship between social support and health. Am J Epidemiol. 1983 May;117(5):521–37.

6. Uchino BN. Social support and health: a review of physiological processes potentially underlying links to disease outcomes. J Behav Med. 2006 Aug;29(4):377–87.

7. Rubino F, Puhl RM, Cummings DE, Eckel RH, Ryan DH, Mechanick JI, et al. Joint international consensus statement for ending stigma of obesity. Nat Med. 2020 Apr;26(4):485–97.

8. Umberson D, Montez JK. Social relationships and health: a flashpoint for health policy. J Health Soc Behav. 2010;51 Suppl:S54-66.

9. Holt-Lunstad J, Smith TB, Layton JB. Social Relationships and Mortality Risk: A Meta-analytic Review. PLOS Med. 2010 Jul 27;7(7):e1000316.

10. Hughes G, Degher D. Coping with a deviant identity. Deviant Behav. 1993 Oct 1;14(4):297–315.

11. Maphis LE, Martz DM, Bergman SS, Curtin LA, Webb RM. Body size dissatisfaction and avoidance behavior: How gender, age, ethnicity, and relative clothing size predict what some won’t try. Body Image. 2013 Jun;10(3):361–8.

12. Rosen JC, Srebnik D, Saltzberg E, Wendt S. Development of a body image avoidance questionnaire. Psychol Assess J Consult Clin Psychol. 1991;3:32–7.

13. Major B, Schmader T. Coping with stigma through psychological disengagement. In: Prejudice: The target’s perspective. San Diego, CA, US: Academic Press; 1998. p. 219–41.

14. Brytek-Matera A, Czepczor-Bernat K, Olejniczak D. Food-related behaviours among individuals with overweight/obesity and normal body weight. Nutr J. 2018 Oct 16;17(1):93.

15. Sarlio-Lahteenkorva S. Relapse stories in obesity. Eur J Public Health. 1998 Sep 1;8(3):203–9.

16. Mehrdad N, Abbasi NH, Nasrabadi AN. The Hurt of Judgment in Excessive Weight Women: A Hermeneutic Study. Glob J Health Sci. 2015 Nov;7(6):263–70.

17. Strauss RS, Pollack HA. Social marginalization of overweight children. Arch Pediatr Adolesc Med. 2003 Aug;157(8):746–52.

18. Karlsson J, Taft C, Rydén A, Sjöström L, Sullivan M. Ten-year trends in health-related quality of life after surgical and conventional treatment for severe obesity: the SOS intervention study. Int J Obes 2005. 2007 Aug;31(8):1248–61.

19. Cohen R, Benvenga R, Fysekidis M, Bendacha Y, Catheline JM. Social isolation but not deprivation involved in employment status after bariatric surgery. PLOS ONE. 2021 Sep 10;16(9):e0256952.

20. Lyons K, Meisner BA, Sockalingam S, Cassin SE. Body Image After Bariatric Surgery: A Qualitative Study. Bariatr Surg Pract Patient Care. 2014 Feb 21;9(1):41–9.

21. Stolzenberger KM, Meaney CA, Marteka P, Korpak S, Morello K. Long-Term Quality of Life Following Bariatric Surgery: A Descriptive Study. Bariatr Surg Pract Patient Care. 2013 Feb 14;8(1):29–38.

22. Coulman KD, MacKichan F, Blazeby JM, Owen-Smith A. Patient experiences of outcomes of bariatric surgery: a systematic review and qualitative synthesis. Obes Rev Off J Int Assoc Study Obes. 2017 May;18(5):547–59.

23. Griauzde DH, Ibrahim AM, Fisher N, Stricklen A, Ross R, Ghaferi AA. Understanding the psychosocial impact of weight loss following bariatric surgery: a qualitative study. BMC Obes. 2018 Dec 3;5(1):38.

24. Chan AW, Tetzlaff JM, Gøtzsche PC, Altman DG, Mann H, Berlin JA, et al. SPIRIT 2013 explanation and elaboration: guidance for protocols of clinical trials. BMJ. 2013 Jan 9;346:e7586.

25. Croy I, Luong A, Triscoli C, Hofmann E, Olausson H, Sailer U. Interpersonal stroking touch is targeted to C tactile afferent activation. Behav Brain Res. 2016 Jan 15;297:37–40.

26. Triscoli C, Croy I, Olausson H, Sailer U. Touch between romantic partners: Being stroked is more pleasant than stroking and decelerates heart rate. Physiol Behav. 2017 Aug 1;177:169–75.

27. Williams KD, Cheung CK, Choi W. Cyberostracism: effects of being ignored over the Internet. J Pers Soc Psychol. 2000 Nov;79(5):748–62.

28. Faul F, Erdfelder E, Lang AG, Buchner A. G*Power 3: a flexible statistical power analysis program for the social, behavioral, and biomedical sciences. Behav Res Methods. 2007 May;39(2):175–91.

29. Pfabigan DM, Frogner ER, Sailer U (2022, December 20). The Social Experiences’ Daily Occurrence Scale (SOLO). Retrieved from osf.io/xcwek

30. Utredning og behandling av sykelig overvekt i spesialisthelsetjenesten Voksne. 2007.https://www.helse-sorost.no/Documents/Styret/Styrem%C3%B8ter/2008/vedlegg-sak-086-2008-Rapport%20-%20utredning%20og%20behandling%20av%20sykelig%20overvekt%20i%20spes%20helsetjenesten%20-%20voksne%20pdf%20211102.Pdf

